# Pulmonary thromboembolism in COVID-19 Patients on CT Pulmonary Angiography - A Single-Centre Retrospective Cohort Study in the United Arab Emirates

**DOI:** 10.1101/2021.11.14.21266214

**Authors:** Ghufran Aref Saeed, Waqar H Gaba, Abd Al Kareem Adi, Reima Al Marshoodi, Safaa Al Mazrouei, Asad R Shah

## Abstract

**Purpose:** Our aim is to identify the prevalence and distribution of pulmonary thromboembolism in COVID-19 infected patients in our hospital.

**Materials and Methods:** Data of all patients with COVID-19 infection either on RT-PCR testing or non-contrast high resolution CT(HRCT) who had CT pulmonary angiography (CTPA) from April to June 2020 were included. 133 patients were initially included in the study, 7 were excluded leaving a total number of 126 patients.

**Results:** Twenty (15.8%) patients had evidence of pulmonary embolism (PE) on CTPA with mean age of 50 years (ranging 31-85) with 95% males. The mean D-dimer was 5.61mcg/mL among the PE-negative and 14.49 mcg/mL in the PE-positive groups respectively. Among the patients with evidence of pulmonary embolism on CTPA almost half required admission to intensive care unit in comparison to only one-fifth with negative CTPA. One-fourth died among the PE positive group with only 5% died among the PE negative group. There was a 33% reduction in the development of PE in the COVID-19 patients who had received low molecular weight heparin (LMWH) prior to their CTPA study versus those who had not.

**Conclusion:** D dimers correlate well with the incidence of pulmonary embolism among COVID-19 patients. Our data suggest that majority of our patients, developed pulmonary embolisms within 5 days into their hospital stay, accounting to almost two thirds of all positive cases diagnosed by CTPA. Those with PE among COVID-19 patients have high chances of ICU admission and mortality. Use of thromboprophylaxis early on might reduce the incidence of PE.

## 1. Introduction

In December 2019, cases of a new virus causing pneumonia have been identified in Wuhan, China [1]. Ever since then, researchers and healthcare workers all around the world have started studying, analyzing, and publishing data on this novel coronavirus. One of the more heavily studied aspects was the virus’s induced prothrombotic state [2] that was strongly associated with increased mortality [3]. Among the notable pathologies caused by the virus’s induced hypercoagulable state was the development of pulmonary emboli. There was an increased prevalence of pulmonary thromboembolism in patients with moderate to severe COVID-19 pneumonia [4]. One meta-analysis reported the incidence of pulmonary embolism in COVID-19 pneumonia patients to be up to 15%, and urged the need to evaluate the roles of both prophylactic and therapeutic anticoagulation [5], as developing pulmonary embolism was shown to increase the morbidity, mortality, prolong the duration of invasive ventilation, and ICU stay [6]. The main objective of our study is to identify the prevalence and distribution of pulmonary thromboembolism in COVID-19 infected patients in Abu Dhabi, UAE; and analyze whether there is a notable difference between the PE positive and PE negative groups regarding their age, gender, CT severity score, D-dimer levels, thromboprophylaxis, and mortality rates.

### 2. Methods

#### 2.1. Data Collection

We obtained ethical approval from the Institutional Review Board (IRB) and Department of Health (DOH), Abu Dhabi, United Arab Emirates (UAE). Waiver of informed consent was allowed by the ethics committee. We collected clinical and laboratory data for analysis derived from an electronic medical record system, from April to June 2020 of patients who were suspected to have COVID-19 infection and underwent a CT pulmonary angiography (CTPA) scan. The results for the CTPA images were collected and evaluated using the Picture Archiving and Communication Systems (PACS).

#### 2.2. CTPA Inspection

All CT Pulmonary Angiograms were performed on the SIEMENS 128 Somatom scanner. Patients were placed in a supine position with a single breath hold. Omnipaque 350 was used as IV contrast with an average volume of 40-50 mL, using a power injector with a flow rate of 4-5 cc/s using a peripheral venous cannula of 18-20 G. Scanning parameters were as follows: topogram length of 512cm, scan direction was craniocaudal, tube voltage of 100 kV and a tube current of 75 mA - smart mA dose modulation. The region-of-interest was the main pulmonary artery with a trigger threshold of 100HU. The monitoring delay was 3 sec with a diagnostic delay of 4 sec, slice thickness was 0.6 mm, pitch was 1.9, rotation time was 0.25 s, and scan time was 0.25 s. Sagittal, and Coronal reformats were also obtained with additional maximum intensity projection images.

#### 2.3. CTPA Image Analysis

The images were evaluated and reviewed by two radiologists with more than 8 years of experience to judge the presence of typical COVID-19 pneumonia findings (bilateral, multilobed, posterior peripheral ground-glass opacities) and judge upon the severity of the infection radiologically.

#### 2.4. Statistical Analysis

Data was analyzed using SPSS 21.0. Demographic, clinical and laboratory data was reported as numbers where each value pertained to a particular subgroup based on their findings (1 for male, 2 for female, etc.). We used the Pearson correlation coefficient for correlations and regarded a p-value less than 0.05 as statistically significant.

#### 2.5. Inclusion criteria consisted of patients having either a positive COVID-19 RT PCR nasal swab or showing typical clinical and radiological pictures highly suggestive of COVID-19 pneumonia

Excluded were patients with an underlying malignancy, and those with a non-diagnostic opacification of the main pulmonary artery on CTPA.

#### 2.6. Thromboprophylaxis

We identified the patients who received low molecular weight heparin (LMWH) prior to their CTPA amongst everyone included in the study, then compared the incidence of PE in them against those who had not. The decision on whether to give thromboprophylaxis, along with the dose of LMWH was decided by the Consultant Physician, guided by a hospital locally approved algorithm (see Appendix A).

## 3. Results

A total of 133 patients underwent CT Pulmonary Angiography during the 3 months period of the COVID-19 pandemic from April to June 2020 at our institution. 126 of those patients were included for the final analysis. 7 were excluded as their COVID-19 PCR was negative and they had no radiological evidence of COVID-19 pneumonia, or they had inconclusive CT scans suggesting the likely presence of a malignancy.

The mean age was 51 years (ranging 29-85). 103 (82%) of patients were males, while the remaining 23 (18%) were females.

Twenty (15.8%) patients had evidence of pulmonary embolism (PE) on CTPA.

Most pulmonary emboli [16/20 (80%)] were in the lobar and distal branches and only 4/20 (20%) were in the main pulmonary arteries (see Appendix B). Among those patients with PE, mean age was 50 years (ranging 31-85). As for their genders, 19 (95%) were male, and only 1 (5%) was female. 34 (26.9 %) CTPA were done upon presentation to the emergency department with a positive incidence of 6/34 (17.6%). 39 (30.9%) CTPA were done 1-5 days into admission with a positive incidence of 7/39 (17.9%), and 53 (42%) CTPA were done 6+ days into admission with a positive incidence of 7/53 (13.2%). (See Appendix C)

It was also evident that almost half (45%) of patients with evidence of pulmonary embolism on CTPA required admission to the intensive care unit during their hospital stay, while only one-fifth (20%) of the 106 patients with no evidence of pulmonary embolism on CTPA did.

The mean D-dimer was 5.61mcg/mL among the PE-negative and 14.49 mcg/mL in the PE-positive groups respectively.

As for mortality rates, there were a total of ten deaths out of all patients studied, with 5% mortality rate in the PE-negative group compared to the 25% mortality rate among the PE-positive group. There was no significant difference in the severity of parenchymal lung involvement radiologically amongst the groups, with most cases being of either the moderate or severe categories almost equally.

Regarding thromboprophylaxis; 83 patients had LMWH prior to CTPA and incidence of PE among those was 12 (14%). On the other hand, 43 patients had not received LMWH or any other type of anticoagulation, and 9 (21%) of those patients had evidence of PE on their scans. As for the dosing, 29(35%) patients received prophylactic dose, 22(26%) received intermediate dose, and 32(38%) received therapeutic dose with incidence of PE being 14%, 13% and 6% respectively (See Appendix D). All doses were adjusted according to patients’ kidney function (creatinine clearance).

## 4. Discussion

Numerous studies of the COVID-19 pandemic have advised early caution and testing against patients developing coagulopathies and venous thromboembolisms including PE [7,8]. It has proven to be a significant burden over the morbidity and mortality rates, reaching mortality rate increases of up to 45% when compared to other general causes [9]. In our study, that risk is evident, and what we would like to highlight in our results is that the majority of our patients who developed pulmonary embolisms did so early on, 0 to 5 days into their hospital stay, accounting to almost two thirds of all positive cases diagnosed by CTPA. That emphasizes the need to be on guard and look for it as a cause early on once patients show any signs of clinical deterioration or have any symptoms suggesting a pulmonary embolism during their hospital stay. We believe early diagnosis and treatment initiation against pulmonary emboli would potentially lower both morbidity and mortality rates, since the subjects studied showed that those who developed pulmonary emboli had substantially higher ICU admission rates and are at a higher risk of death. We also suggest that, like other studies have shown regarding the issue, D-dimer acts as a decent indicator of hypercoagulability [10,11], given that all our patients who developed pulmonary embolisms on CTPA had positive D-dimers, and their mean D-dimer level was much higher than that of the PE-negative group.

Even though it’s been shown through evidence that increasing CT severity score did to an extent predict patient’s clinical status, particularly their oxygen requirement [12], we could not demonstrate that it had any predictive value on patients developing pulmonary embolism with a moderate to severe chest HRCT score. [See Appendix E for more information on how HRCT severity scoring was calculated]

Overall, acute pulmonary embolism developed in 20/126 (15.8%) of the cases studied over the 3-month period. which is more or less consistent with the currently available worldwide statistic [5]. Moreover, regarding thromboprophylaxis; when the effect of receiving LMWH prior to the patients’ CTPA was analyzed, even though both groups are not identical in number and lack information regarding certain control parameters (such as severity and duration of illness) there was a 33% reduction in the development of PE in the COVID-19 patients who had received LMWH prior to their CTPA study versus those who had not.

## 5. Conclusion

In patients diagnosed with COVID-19 infection either by RT-PCR or high clinical and radiological suspicion, we should be aware of this evident risk of hypercoagulability and embolism to the lung’s vasculature, whose symptoms could be masked as a mere deterioration of the patient’s respiratory status. Being vigilant in diagnosing and treating such incidents could potentially reduce morbidity and mortality rates of the COVID-19 pandemic. Our study also shows benefit in giving thromboprophylaxis (LMWH at prophylactic doses or higher), leading to a potential decrease in the incidence of PE among such patients.

**Figure 1:**
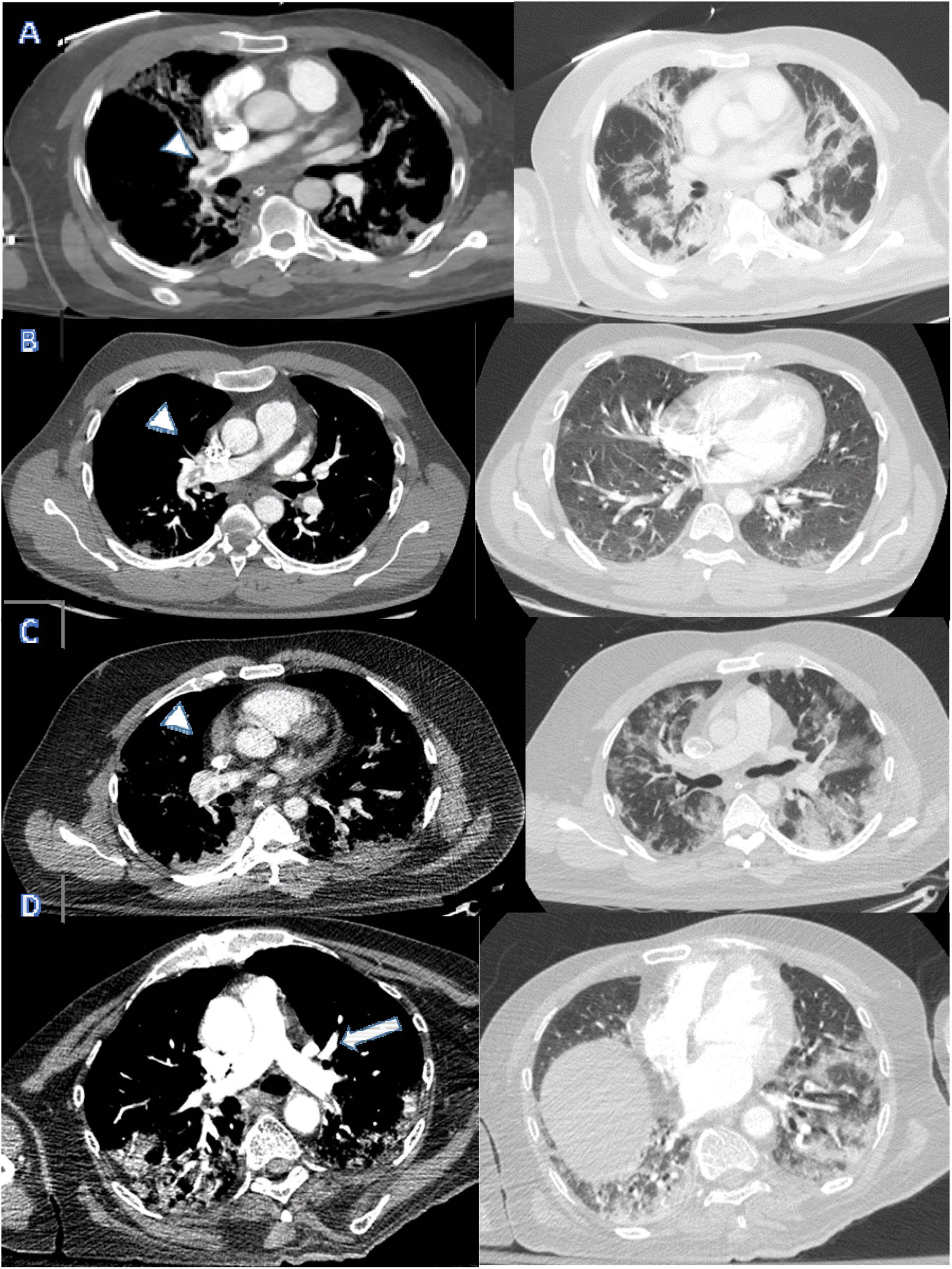
Four Cases of Central Pulmonary Embolism presenting to the ED. Axial soft tissue window (left pane) showing right main pulmonary artery emboli (Arrow-heads in A-C) and small proximal left lower lobar pulmonary artery embolus (Arrow in D). Corresponding lung windows demonstrating the typical sub-pleural Covid pneumonia changes.

**Figure 2:**
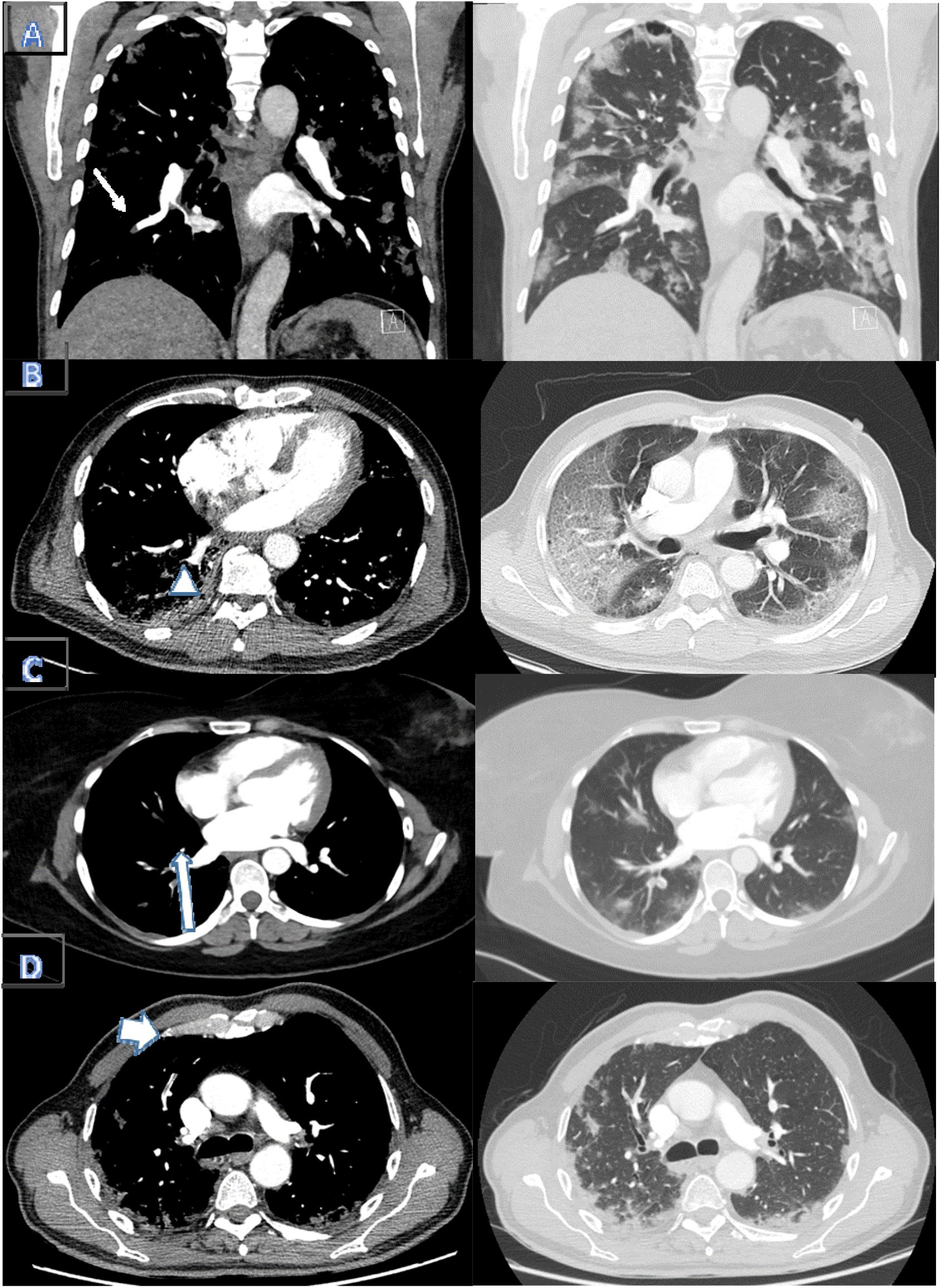
Axial soft tissue window images (left pane) showing segmental emboli in the right lower lobe (Thin arrow in A, Arrow head in B and Long arrow in C) and right upper lobe (Broad arrow in D). Corresponding lung window images showing typical changes of Covid pneumonia.

**Table 1:** Individual lobar scores based on percentage of involvement.

| Lobar involvement | Score |
| --- | --- |
| 5% or less | 1 |
| 5%–25% | 2 |
| 26%–49% | 3 |
| 50%–75% | 4 |
| >75% | 5 |

**Table 2:**
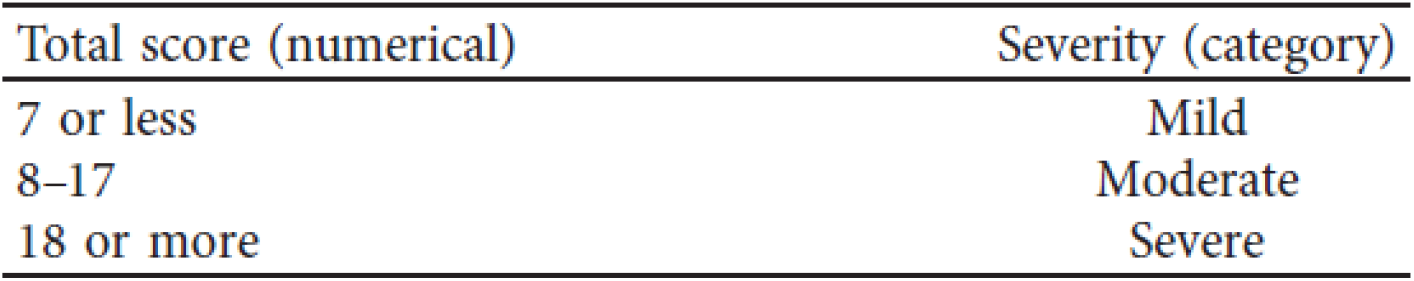
Sum of the individual lobar scores indicates the overall severity of the five lobes.

| Total score (numerical) | Severity (category) |
| --- | --- |
| 7 or less | Mild |
| 8–17 | Moderate |
| 18 or more | Severe |

## Data Availability

Data are available on request through Institutional Review Board

**Appendix A:**
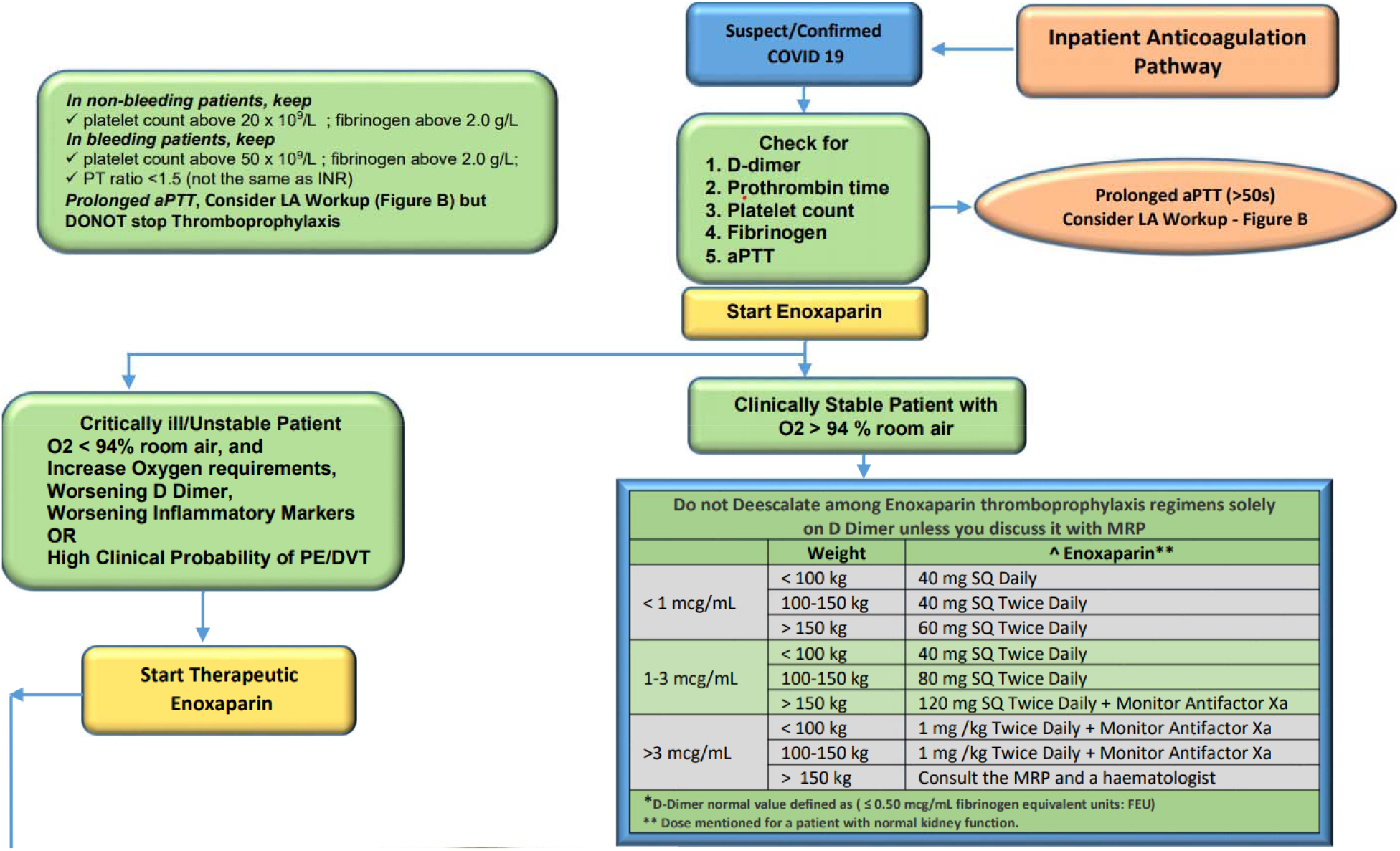
Algorithm used to guide physicians to choose LMWH dosing.

**Appendix B:**
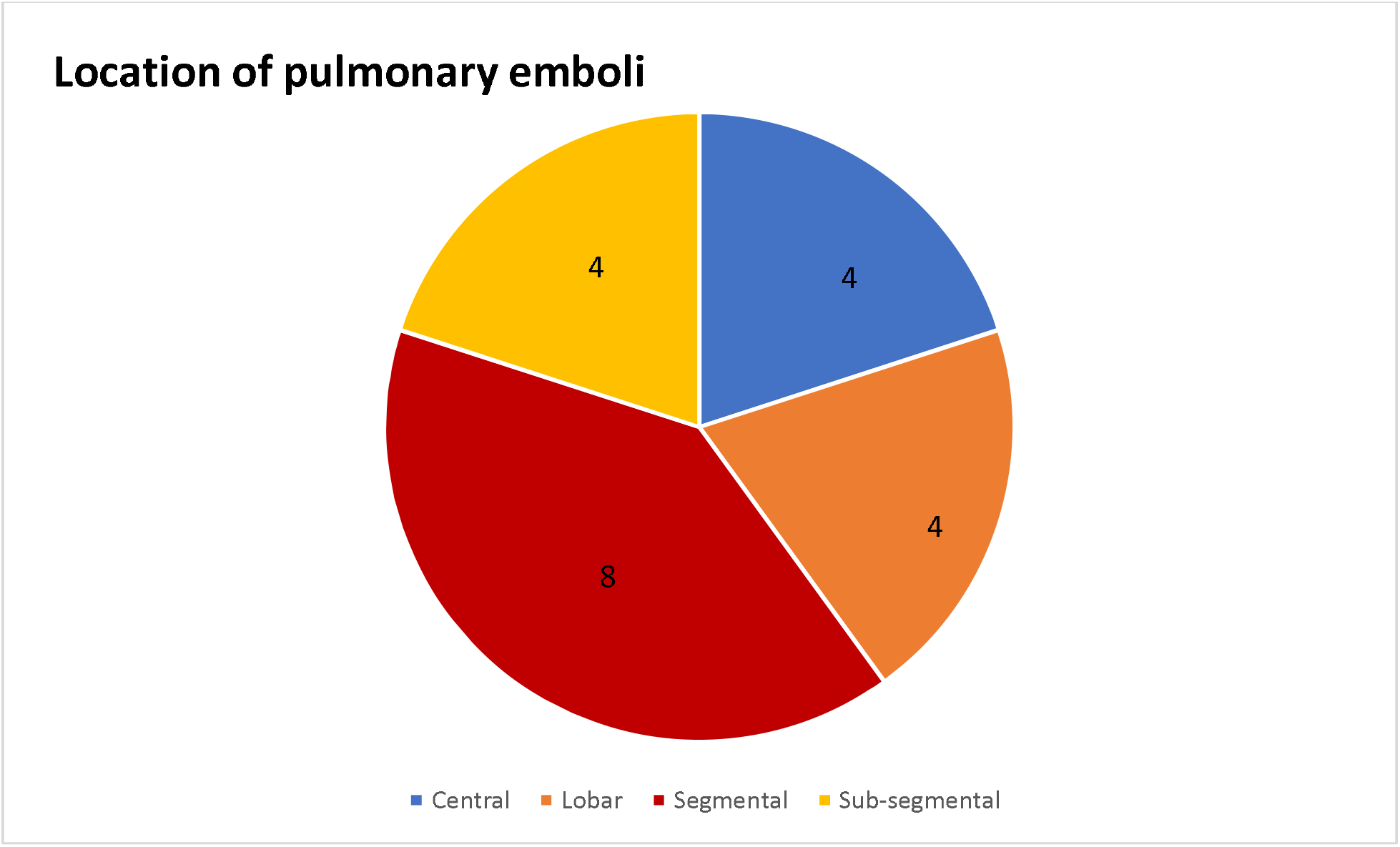
General location of pulmonary emboli.

**Appendix C:**
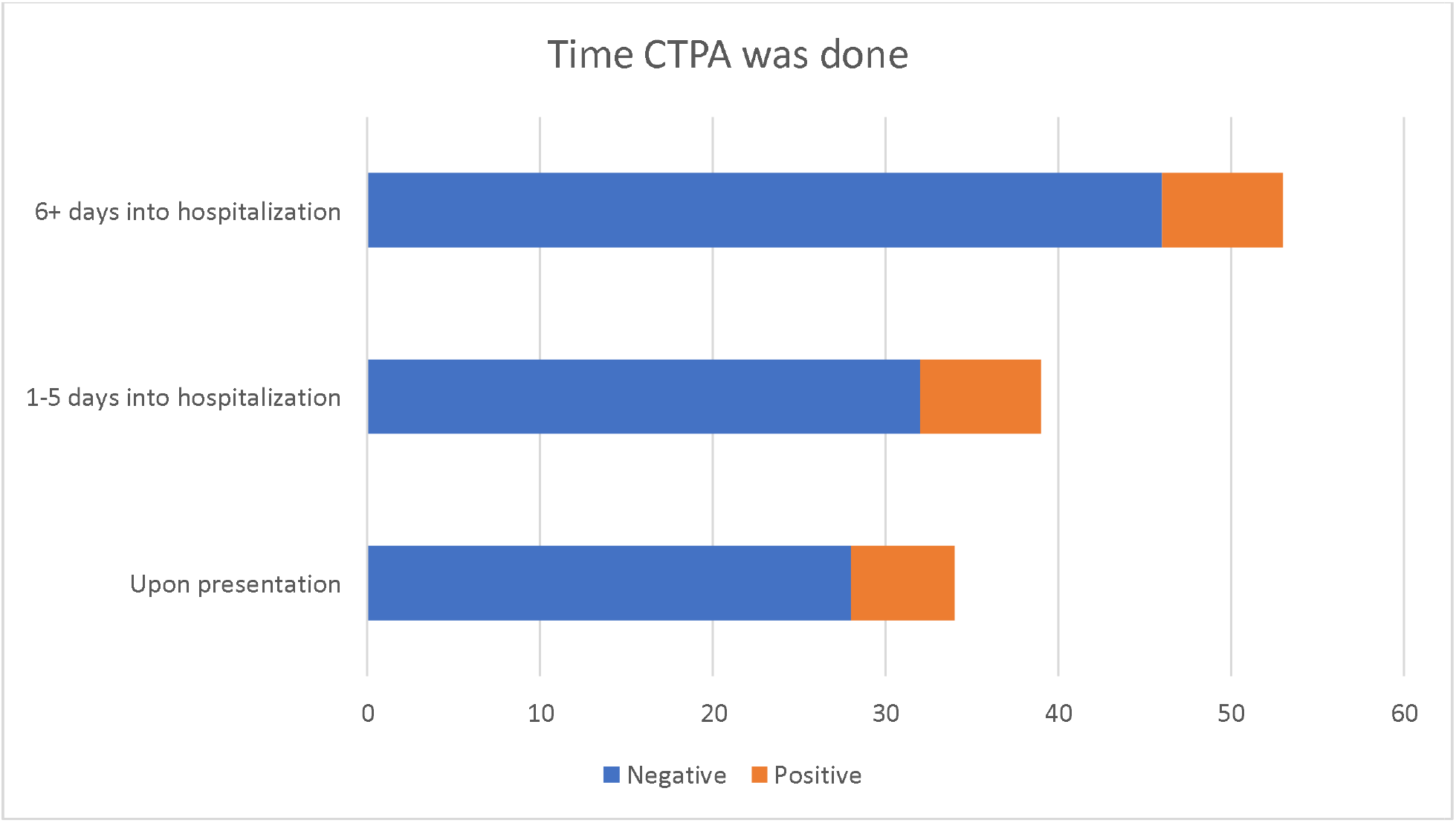
Time CTPA was done during hospitalization

**Appendix D:**
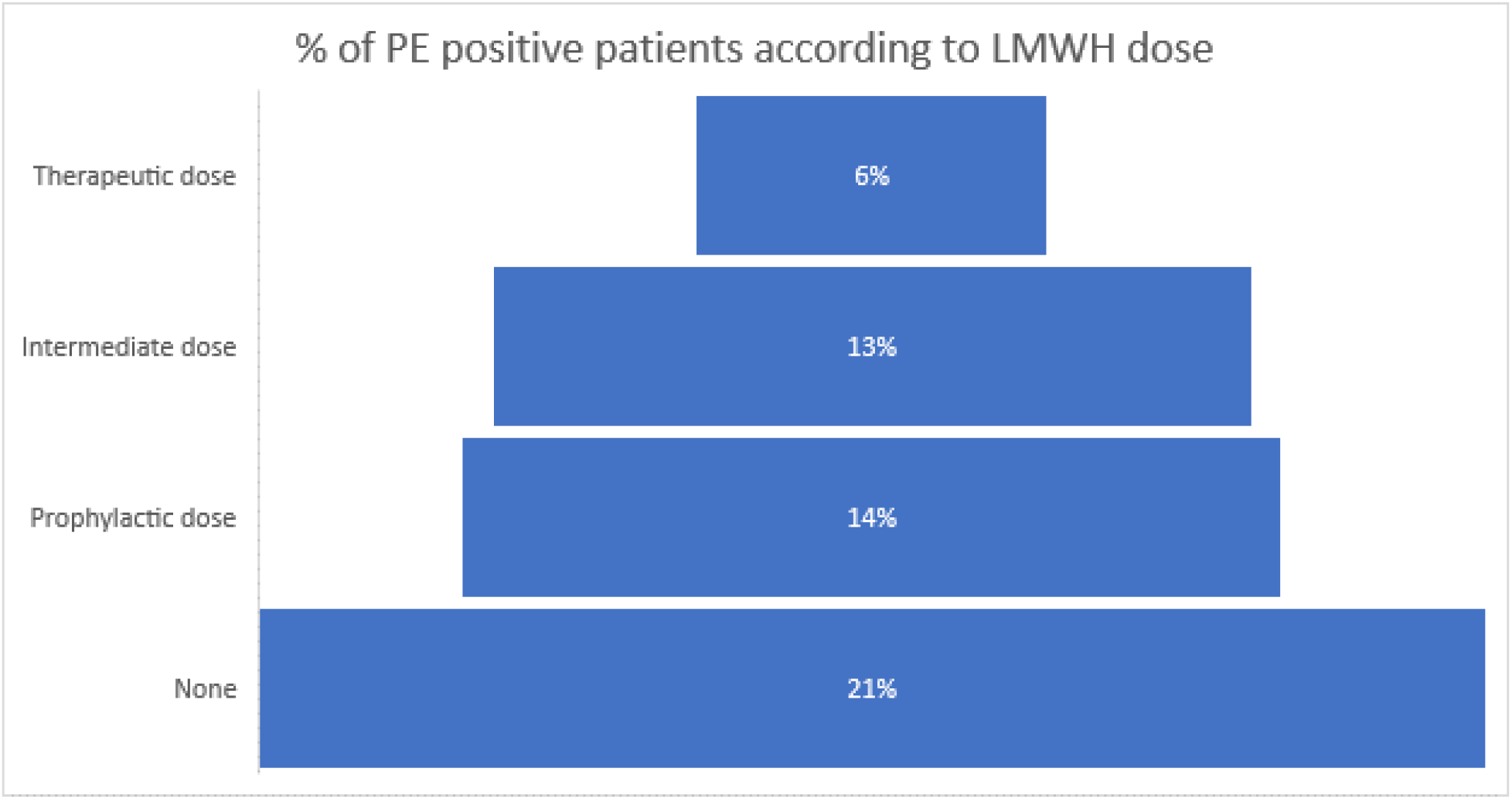
Percentage of PE positive patients in the subgroups classifying dose of LMWH received.

## Appendix E

Table used to calculate severity score based on HRCT chest analysis. Each lung lobe was given a score of 1-5 based on degree of parenchymal involvement, and the total was then calculated to categorize severity. [12]

## Abbreviations

COVID-19: Coronavirus disease CT Computed Tomography
CTPA: Computed tomography pulmonary angiography HRCT High-resolution computed tomography
RT-PCR: Reverse transcriptase polymerase chain reaction LMWH Low molecular weight heparin
PE: Pulmonary embolism
ICU: Intensive care unit
PACS: Picture archiving and communication systems

## Data Availability

Data are available on request through Institutional Review Board

## Acknowledgements

The authors would like to thank CT radiographers and technical department of Radiology department of Shiekh Khalifa Medical City, Abu Dhabi.

